# Controlling the Spread of COVID-19: Optimal Control Analysis

**DOI:** 10.1101/2020.06.08.20125393

**Authors:** Chinwendu E. Madubueze, Sambo Dachollom, Isaac Obiajulu Onwubuya

## Abstract

Coronavirus disease 2019 (COVID-19) is a disease caused by Severe acute respiratory syndrome coronavirus 2 (SARS CoV-2). It was declared on March 11, 2020, by the World Health Organization as pandemic disease. The disease has neither approved medicine nor vaccine and has made government and scholars search for drastic measures in combating the pandemic. Regrettably, the spread of the virus and mortality due to COVID-19 has continued to increase daily. Hence, it is imperative to control the spread of the disease particularly using non-pharmacological strategies such as quarantine, isolation and public health education. This work studied the effect of these different control strategies as time-dependent interventions using mathematical modeling and optimal control approach to ascertain their contributions in the dynamic transmission of COVID-19. The model was proven to have an invariant region and was well-posed. The basic reproduction number was computed with and without interventions and was used to carry out the sensitivity analysis that identified the critical parameters contributing to the spread of COVID-19. The optimal control analysis was carried out using the Pontryagin’s maximum principle to figure out the optimal strategy necessary to curtail the disease. The findings of the optimal control analysis and numerical simulations revealed that time-dependent interventions reduced the number of exposed and infected individuals compared to time-independent interventions. These interventions were time-bound and best implemented within the first 100 days of the outbreak. Again, the combined implementation of only two of these interventions produced a good result in reducing infection in the population, while the combined implementation of all three interventions performed better, even though zero infection was not achieved in the population. This implied that multiple interventions need to be deployed early in order to the virus to the barest minimum.

## Introduction

The Novel Coronavirus, severe acute respiratory syndrome coronavirus 2 (SARS-CoV-2) is a new strain of coronaviruses that cause the coronavirus disease 2019 (COVID-19) and was declared a pandemic by the World Health Organization (WHO) on 11^th^ March 2020 [1]. The virus was discovered in December 2019 in Wuhan City of Hubei Province, China [2, 3]. They belong to the order of Nidovirales, a family of Coronaviridae, and subfamily of Orthocoronavirinae [4]. Coronaviruses are a group of enveloped viruses with a positive-sense, single-stranded RNA and viral particles resembling a crown from which the name was derived [3].

The COVID-19 is a highly infectious disease that can be spread directly or indirectly from an infectious person to a healthy person through the eye, nose, mouth, and sometimes through the ears via droplets produced when coughing or sneezing [2, 5, 6]. The exact source of the disease is uncertain. However, rodents and bats have been suspected by many researchers [5, 7]. The SARS-CoV-2 can survive up to 8-10 hours over porous surfaces (like paper, wood, sponge and fabric etc) and a little more than 8-10 hours on non-permeable surfaces (glass, plastics, metals etc) [2]. It has an incubation period of usually 2 – 14 days [2, 8]. Its symptoms are similar to that of common cold or flu. Also, others include fever, dry cough, shortness of breath and pneumonia [5]. The severity of the illness can vary in different people from mild to severe symptoms based on their age and health status [2, 5, 6]. Almost 80% of COVID-19 patients are either asymptomatic or have mild symptoms and usually recover from the disease within 2 weeks. However, high mortality is recorded among the aged people and people with underlying chronic diseases, 2% of COVID-19 sufferers were usually under 18 years of age, out of which, fewer than 3% developed severe conditions [2]. COVID-19 has a low mortality rate that ranges from 2-3% which is significantly less than 10% of the severe acute respiratory syndrome (SARS) in 2003 and 35% of Middle-East respiratory syndrome (MERS) in 2012 [2, 5, 9, 3]. Due to its high infectivity, COVID-19 spread exponentially to virtually every part of the world within three months [1, 10].

As of April 2020, almost every country of the world has recorded at least one positive case despite speculation that the virus doesn’t thrive in regions with hot weather. Entire Europe (especially, Italy and Spain) has become the epicenter of the outbreak, while the United States of America, Asia, and Australia record hundreds of new infections daily, with thousands of disease mortalities recorded [10]. In Africa, Nigeria recorded her first case on February 27, 2020, but as of April 26, 2020, just within 60 days, the figure has risen to 1364 infections with 45 deaths so far [11]. It is sad to note that most of the mortalities of COVID-19 especially in developing countries are attributed to poor medical facilities and manpower.

There is no specific treatment or vaccine available for COVID-19 probably because it is a new disease and vaccine development usually take up to 18 months [9, 12]. There is no approved medicine that eradicates the virus, however, treatment is mainly supportive [2, 9, 13]. It is because of these realities that governments across the world have resorted to non-pharmacologic measures. For instance, the Nigerian government has sensitized its citizenry on the need to adopt safety measures such as wearing of disposable surgical face masks, regular hand-washing with plenty of soap under running water, and the use of alcohol-based hand sanitizer in the absence of soap and water among others as recommended by the WHO [1, 10, 11]. Also, many governments worldwide are spending billions of the United States’ dollars as well as soliciting aids from well-spirited individuals and organizations towards combating the COVID-19 pandemic. Furthermore, many countries have imposed compulsory self-quarantine and restricted movements of their citizenries (lockdown/sit at home), closure of businesses, and borders as preventive measures [1, 10]. These interventions have succeeded greatly in curtailing the trans-border spread of the SARS-CoV-2 from country to country. Nevertheless, the emerging major problem in the spread of COVID-19 is human-to-human transmission in a heterogeneous community. Sadly, the implementation of these interventional policies of governments (e.g. total lockdown of movement, businesses, fear of quarantine/isolation), has thrown up another new challenge in the fight of the disease because of hunger and poverty especially in developing countries in sub-Saharan Africa where governments lack social securities. Therefore, there is the need to find cost-effective ways of halting the COVID-19 pandemic with minimal economic and social disruptions to avert impending catastrophic economic rupture.

Scholars are approaching this pursuit from two broad but complementary aspects of sciences; the Medical sciences and natural sciences. The medical scientists are busy trying to identify the source(s) of the disease, quicker ways of detecting the disease, treatment, and vaccine production [1, 12, 13]. The natural scientists are busy trying to proffer interventional measures through the development of mathematical models that will control the disease transmission especially now that there is no vaccine or known treatment.

Mathematical models have over the years proven to be reliable and efficient tools employed in formulating control strategies towards suppressing and mitigating the effects of infectious diseases, epidemics and pandemics such as Ebola, SARS, MERS etc [14, 15, 16]. For COVID-19, some mathematical models have been produced which aim at halting the spread of the disease and forecasting its transmission through simulations. Some scholars focused on calculating the basic reproduction number, (*R*_0_)[8, 17, 18, 19] and failed to consider the effect of public health education, quarantine and isolation on the transmission of COVID-19. In the work of Imai *et al*. [9], they assumed COVID-19 is highly inconsistent in terms of the number of new infections just like SARS. They also investigated the consistency of their model with realities of the outbreak size using the set of simulated epidemic paths. Their findings affirm that without the implementation of holistic control measures, human-to-human transmissibility of COVID-19 is enough to sustain the pandemic and postulated that, COVID-19 will have a diminutive generation time if the majority of COVID-19 cases are mild to moderate symptoms. Shen *et al*. [20] credited the high case detection rate and quick response by China and the world at large, to experiences from fighting the previous coronaviruses. Their findings postulate that COVID-19 may be a weak specie in the coronavirus family, using the national epidemic of Wuhan in China with a fatality rate of 11.02% (9.26-12.78%) which is less than to those of SARS (14-15%) and MERS (34.4%) with a total of 8042 (95%CI: 4199-11884) infections and 898 (368-1429) death respectively. Chen *et al*. in their research simulated the potency of transmission of COVID-19 from bat (probable) source to humans using their Bats-Hosts-Reservoir-People transmission model [6]. They calculated the basic reproduction number (*R*_0_)using the next generation matrix approach and their results revealed that COVID-19 has higher transmissibility than MERs in the Middle East countries. Rabajante [21] reveals that more havoc and transmission/spread of COVID-19 is being perpetrated within the period an infected person is exposed. Rabajante [21] stated that such an infected person can transmit the virus, especially in a social/public gathering in a remote community within 14 days infectious period. Using the early models of COVID-19, Rabajante [21] recommends the maximum observance of control measures in any public/social events. Tang *et al*. [3] updated their previous model to a time-dependent model. They took into cognizance new interventional advances made in the COVID-19 fight. Their updated findings reported that the best control measure is persistent and constant strict self-isolation. They predicted that the pandemic will peak if the public health measures are adhered to. He *et al*. [6] studied the transmission of COVID-19 with binomial distributions in their discrete-time stochastic epidemic model. Their model parameters were derived from fitted reported data of China from January 11 to February 13, 2020. Their basic reproduction number affirms the positive contributions of various control measures recommended by WHO. While the result of numerical simulation suggests that the disease will peak around February 2020 with contact rate as a paramount factor in the control of COVID-19.

Bordered on how best to control the disease with minimal risk (optimization theory), assessing the consequences of some interventional measures and the risk involved especially now that antiviral treatment and vaccines are not yet available, it is crucial to investigate the optimal control of some control measures. Optimal control is the generalization of the classical calculus of variation in optimization theory. It involves minimizing the cost function and converting a given optimal control model into a Hamiltonian function and apply the Pontryagin’s maximum principle. Optimal control has been successfully applied to infectious diseases like HIV, Ebola, Tuberculosis, and SARS. For a disease like COVID-19 that spread fast, the timing of implementing control measures are important. Unfortunately, very few researchers like [22, 23] considered an optimal control analysis of the COVID-19 transmission and suggest that more researches should be directed in this regard. Djidjou-Demasse *et al*. [22] in their work employed the concept of optimal control theory to explore the best control strategy to implement while awaiting the vaccine. They deduce that the only end to COVID-19 is when humans develop immunity. They weigh the options of humans developing natural immunity after infection or after been vaccinated. Their findings reveal that vaccination will best minimize the cost of loss of human lives, while maximal implementation of control strategies will peak the pandemic in four months after onset. Their results forecasted the possibility of having an efficient vaccine to be in 18 months. Moore and Okyere [23] attributed the rapid spread of COVID-19 to poor medical amenities. Their optimal control analysis focused on the controls; personal protection, treatment and environmental spraying (environmental hygiene) as time-dependent control functions. Their numerical simulation reveals that optimal implementation of all the control measures greatly reduces exposed and infectious individuals in the population.

From the foregoing literature, interventions have been invested in, advocated for and implemented by various stakeholders and still ongoing in the fight of COVID-19. These have cost a huge sum of money and time, casualties in businesses, economic, lives etc. Unfortunately, the world is still recording high mortality and morbidity due to the disease. The few mathematical models that abound on COVID-19 suggesting diverse interventional control measures are yet to explore critically the optimal control analysis of those control parameters. This is necessary to ascertain their contributions in the dynamic transmission of COVID-19 for guidance in formulating better policies on the fight against COVID-19.

The model by Gumel *et al*. [16] forms the motivation for this study. Gumel *et al*. studied the impact of quarantine and isolation on the transmission dynamics of SARS. They assumed that everyone quarantined progress to isolation. The control measures in their work were assumed to be time-independent control measures. But there is a possibility that some people will not develop symptoms after the quarantine. So they return to susceptible class to avoid being infected in the isolation centre. Also, it is well known that behavioral change played a very important role in the spread of disease. Public health education contributed to people’s behavioral changes towards infectious diseases such as Cholera [24], and Ebola virus disease [25]. It will help the health personnel to reach out to people and influence them to adopt new behavioral changes and practice personal hygiene. Thus, the study seeks to ascertain the effectiveness of public health education, quarantine and isolation in reducing the infection of COVID-19 and the time taken to achieve that. It will establish the optimal control strategies required and the proportion of exposed individuals that will be quarantine to curtail the disease. It will seek the effect of time-dependent control variables and control constants on the transmission dynamics of COVID-19. The effect of constant controls will be explored using sensitivity analysis which will be used to identify the most sensitive model parameter that will be targeted. Pontryagin’s maximum principle will be applied to the optimal control model.

The rest of the paper is organized as follows: Section 2 is the model formulation for the COVID-19 with control measures. The model analysis for the COVID-19 is discussed in Section 3 with a sensitivity analysis of the model parameters. The optimal control of the COVID-19 model and its analysis are described in section 4 while Section 5 is the numerical simulations and its discussion. Section 6 is the conclusion.

### Model Formulation

In this section, the formulation of a deterministic model for COVID-19 is presented. The model by Gumel *et al*. [16] used for the control of the SARS outbreak is extended for the control of COVID-19 in this study. The total population, *N*(*t*), at time, *t*, is divided into subpopulations/compartments; Susceptible, *S*(*t*), Exposed, *E*(*t*), Quarantined, *Q*(*t*), Infectious not hospitalized, *I*(*t*), Hospitalized/Isolated Infectious, *H*(*t*), and Recovered, *R*(*t*). We further extend their model by incorporating public health awareness and the possibility of persons in the Quarantined, *Q*(*t*) who tested negative for COVID-19 to return to the Susceptible, *S*(*t*). The quarantined compartment comprised persons who came from high-risk regions and contacts of those who tested positive for COVID-19. These persons are kept for the incubation period of the virus. They are tested for the virus within this period. Those who tested negative returned to Susceptible, *S*(*t*)while those who tested positive are taken to the compartment of Hospitalized/Isolated Infectious, *H*(*t*). Those who missed quarantine but tested positive are in the Infectious not hospitalized, *I*(*t*)compartment from where they either recover or enter the compartment of Hospitalized/Isolated Infectious.

The human population at any given time, *t*, is given by

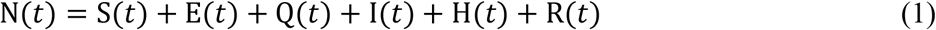

In the Susceptible compartment, *S*(*t*), a proportion (1 *− π*) of humans are recruited into the population at a constant rate, *Λ*, through immigration/emigration of no risk population and through a proportion, *q*, of quarantine individuals that returned to susceptible compartment after 14 days without symptoms at the rate, *σ*_1_. People exit the susceptible compartment either through infection induced by the disease with the force of infection, Ψ. The awareness campaign, α(*t*) ∈ [0,1], reduces the force of infection, Ψ and it is time-dependent. This yields

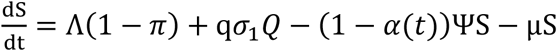

where

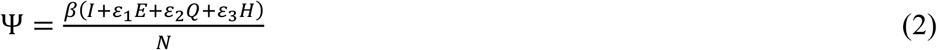

and the parameters, *ε*_1_ *ε*_2_, *ε*_3_, are the modification factors for the exposed, quarantine and hospitalized/isolated individuals. The parameters, *ε*_2_, *ε*_3_, are associated with the hygiene consciousness of the quarantine and the hospitalized/isolated individuals.

The exposed compartment, *E*(*t*), gains population through infection induced by the disease at the rate of (1 *−* α)ΨS and from a proportion, *π*, of the recruitment of people immigrating from a high-risk population of COVID-19 at the rate of*Λ*. A proportion, *p*, of the exposed individuals, exit through quarantine at the rate *τ*(*t*), and the remaining proportion, (1 *− p*) of the exposed individuals escape the quarantine and progress to the infected compartment at the rate, *ρ*, due to ignorance or fear of been quarantine. This gives

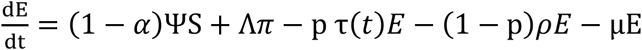

For the quarantined compartment, *Q*(*t*), it gains population from the proportion,*p*, of the exposed human at the rate, *τ*(*t*). A proportion, *q*, of the quarantined individuals exits back to susceptible class after 14 days of no symptoms and reexamination at the rate, *σ*_1_ while a proportion, (1 *− q*) of the quarantined individuals that tested positive progress to Hospitalized/Isolated compartment at the rate, *σ*_2_. That is,

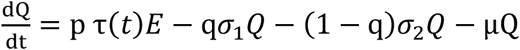

The infectious not hospitalized compartment, *I*(*t*), gains population from a proportion, (1 *−* q), of the exposed individuals that escaped been quarantined and later develop symptoms and progress at the rate, *ρ*. Also, the infectious individuals are isolated at the rate, *η*(*t*). The infectious compartment that escaped isolation can be recovered due to a boost in immunity at the rate of, *γ*_1_, and progress to the recovered compartment or die of the virus at the rate,*d*_1_. This gives

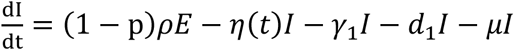

The Isolated/Hospitalized compartment, *H*(*t*), gains population from a proportion, (1 *− q*), of quarantined humans that become infectious during the 14 days quarantine period at the rate, *σ*_2_, and the infectious not hospitalized individuals that are isolated at the rate, *η*(*t*). People exit the hospitalized/isolated compartment through recovery at the rate, *γ*_2_, or death-induced rate, *d*_2_. This yields

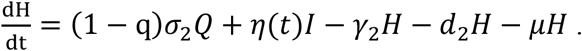

The recovered compartment, *R*, gain population from the infectious not hospitalized individuals that miss isolation but recover due to boost in immunity, and also from the hospitalized/isolated individuals at the rates of *d*_1_ and *d*_2_ respectively. The description of the parameters used in the COVID-19 model is given in Table 1 and the parameters are assumed to be positive.

**TABLE 1:**
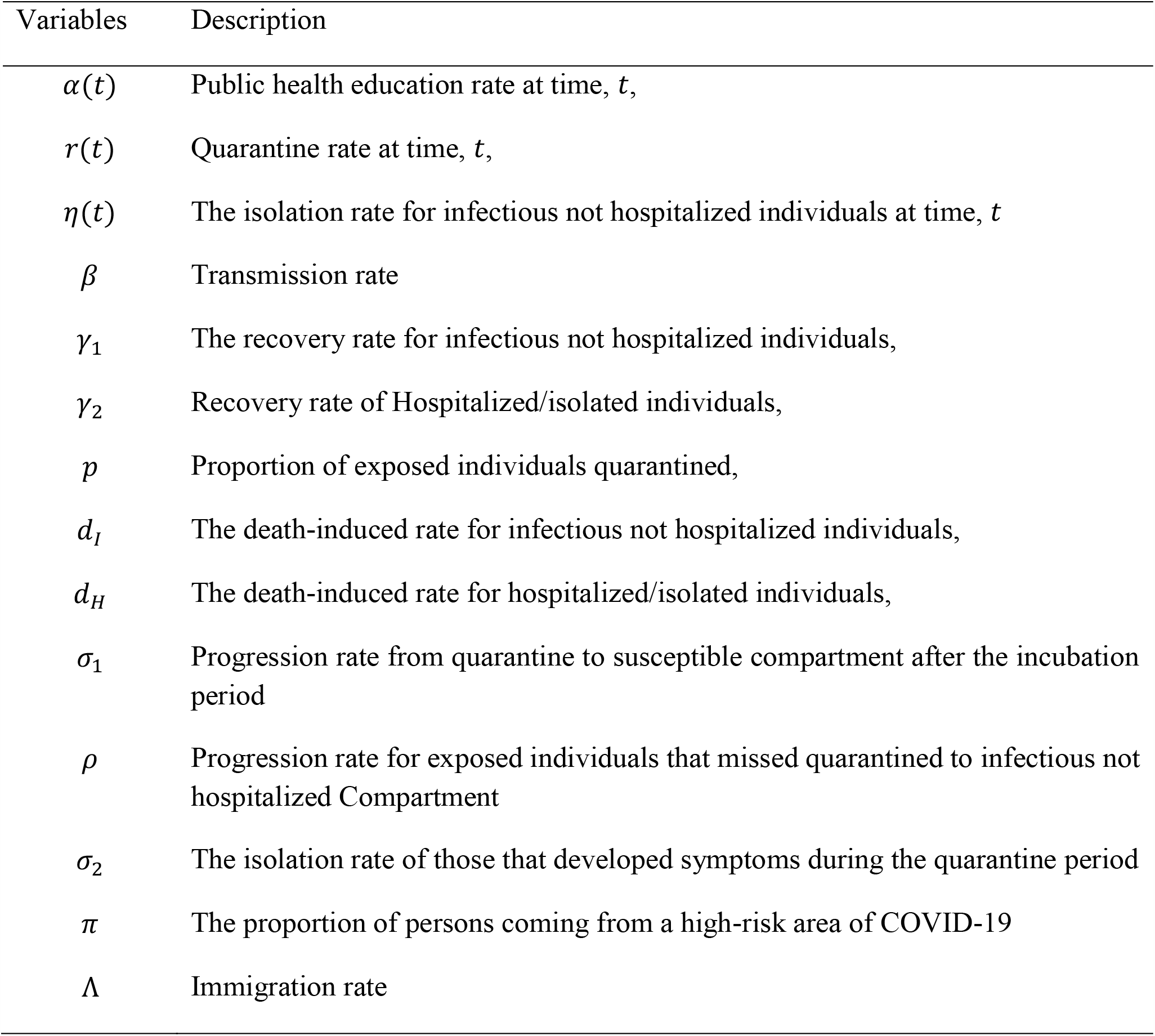
Description of parameters of the Model

| Variables | Description |
| --- | --- |
| $\alpha(t)$ | Public health education rate at time, $t$, |
| $r(t)$ | Quarantine rate at time, $t$, |
| $\eta(t)$ | The isolation rate for infectious not hospitalized individuals at time, $t$ |
| $\beta$ | Transmission rate |
| $\gamma_1$ | The recovery rate for infectious not hospitalized individuals, |
| $\gamma_2$ | Recovery rate of Hospitalized/isolated individuals, |
| $p$ | Proportion of exposed individuals quarantined, |
| $d_I$ | The death-induced rate for infectious not hospitalized individuals, |
| $d_H$ | The death-induced rate for hospitalized/isolated individuals, |
| $\sigma_1$ | Progression rate from quarantine to susceptible compartment after the incubation period |
| $\rho$ | Progression rate for exposed individuals that missed quarantined to infectious not hospitalized Compartment |
| $\sigma_2$ | The isolation rate of those that developed symptoms during the quarantine period |
| $\pi$ | The proportion of persons coming from a high-risk area of COVID-19 |
| $\Lambda$ | Immigration rate |

The system diagram for the transmission of COVID-19 is shown in Figure 1 and the assumptions of the model are given below.

**Figure 1.**
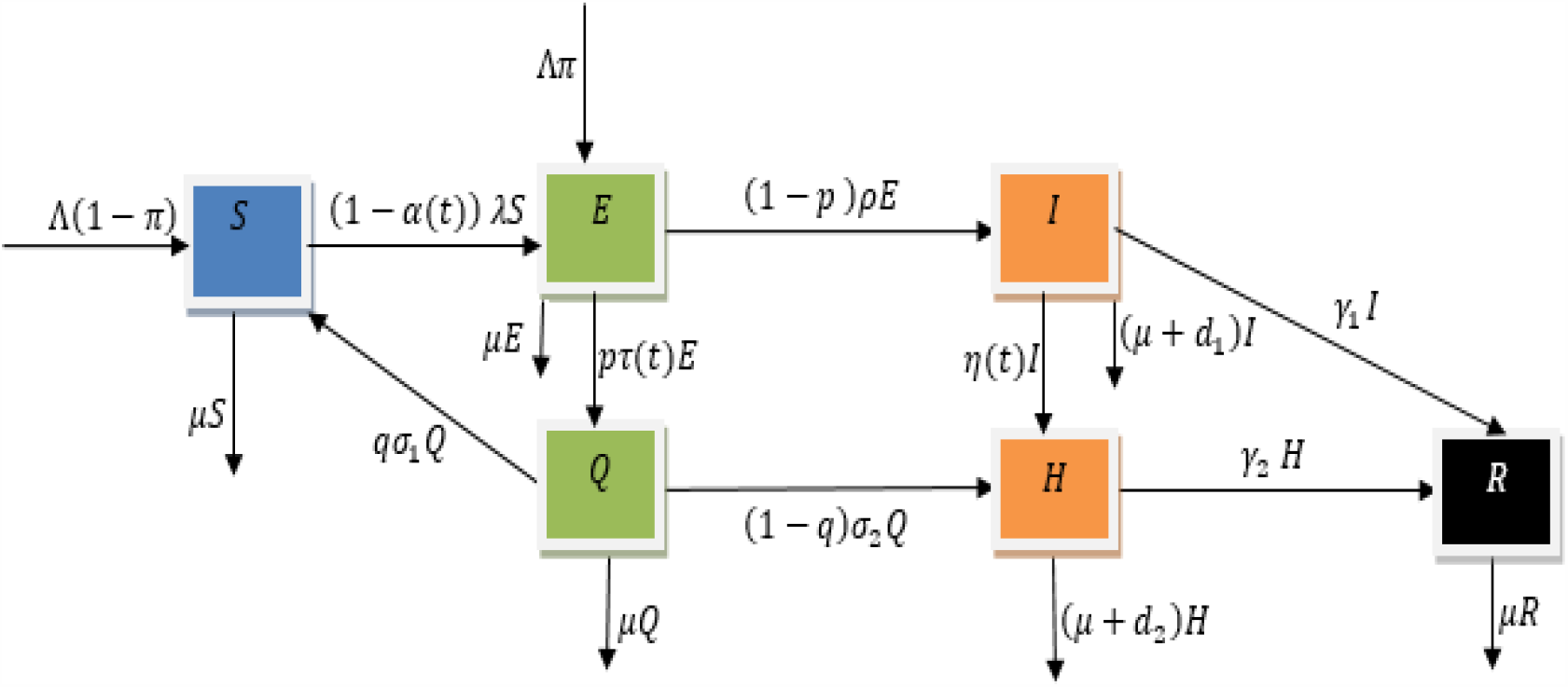
The systematic diagram of the COVID-19 model.

### Model assumptions

i. The recovered individuals are assumed to develop permanent immunity to COVID-19 and all the compartments exit their compartments through natural death rate, *μ*.
ii. There are vital dynamics attributed to the inflow of immigrants, as well as natural deaths. This allows a demographic process to take place.
iii. Infection is acquired via direct contact with infectious human contaminants or droplets and inhalation of infected aerosols only. Rodent infection is not considered due to the reality that present infections that are ravaging the world are mostly secondary infections via humans and humans contaminants.
iv. For an individual to become infectious, he/she must pass through the latent stage.

From the schematic diagram in Figure 1, the model equation is derived

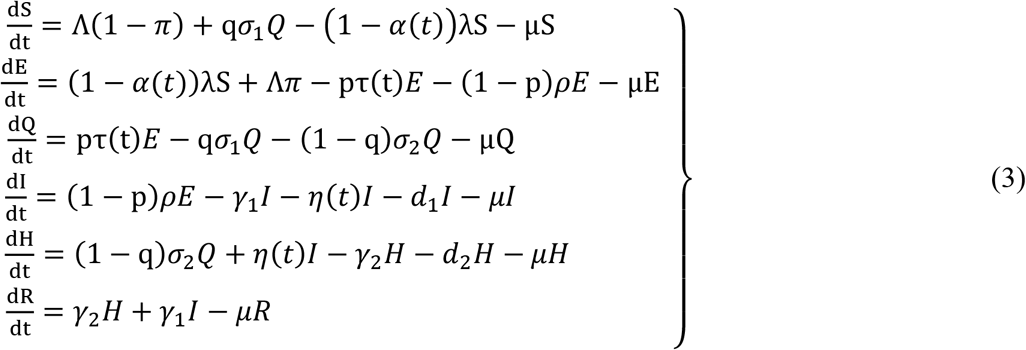

with S(0)> 0, *E*(0)≥ 0, *Q*(0)≥ 0, *I*(0)≥ 0, *H*(0)≥ 0, *R*(0)≥ 0 as the initial conditions.

The control effort, *α*(*t*), represents the public health education effort in educating people about the importance of social distancing, stay at home, and hand-washing in halting the spread of COVID-19. The control effort, *τ*(*t*), represents the effort used to quarantine the exposed individuals and the control effort, *η*(*t*) represents the effort used to isolate the infected individuals. The public health education effort involves educating the public through social media, television, radio, churches, mosques, traditional rulers in the community on how to observe social distancing and hand washing. The efforts used to quarantine the exposed individuals and isolate infected individuals include recruiting and training of the health workers on how to wearing personal protective equipment (PPE), tracing the contacts of those exposed to the COVID-19 through home visits and phone calls, counseling, provision of an ambulance to convey the infected individuals to the isolation centre, general/ COVID-19 tests, provision of isolation centres for treatment and other related logistics.

Our goal is to minimize the cost function given as

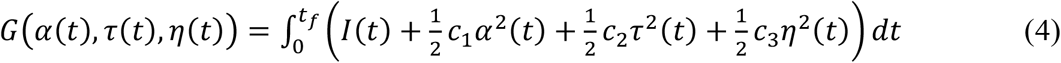

subject to the system of differential equations (3). All control efforts, *α*(*t*), *τ*(*t*), *η*(*t*), are assumed to be bounded and Lebesgue measurable time-dependent functions on the interval [0, *t*_*f*_], where *t*_*f*_ is the final time. The control effort set is defined as

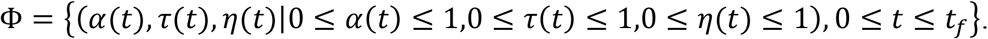

The parameters, c_1_, c_2_ and c_3_, are the balancing cost factors for the public health education effort, *α*(*t*), the quarantine efforts, *τ*(*t*) and the isolation effort, *η*(*t*) respectively. The terms, 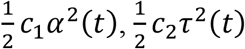 and 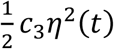 represents the costs associated with public health education, quarantine and isolation respectively. Based on the literature for the optimal control of epidemics, the cost of the controls is assumed to be nonlinear and quadratic [26, 27]. If *α*(*t*)= *τ*(*t*) = *η*(*t*) = 1, then 100% effort is applied in public health education, quarantine and isolation respectively at time, *t*. Conversely, if *α*(*t*) = *τ*(*t*) = *η*(*t*) = 0 then no public health education for the people, no quarantine is carried out for the exposed (latent) individuals and no isolation for the infected not hospitalized individuals.

### Model Analysis

For the sake of model analysis, the controls, *α*(*t*), *τ*(*t*), *η*(*t*)are considered as constants, that is, *α*(*t*)= *α, τ*(*t*)= *τ, η*(*t*) = *η*. Some properties of model analysis will be carried out to understand the impact of the constant control parameters on the transmission dynamics of the COVID-19.

### Invariant region

The solutions of the model are uniformly bounded in a positive invariant region,

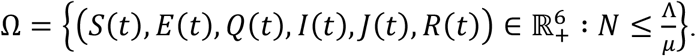

The total population at any time, *t*, is given by (1) and 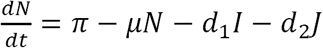.

When the disease-related death rates, *d*_1_, *d*_2_, are neglected, we have

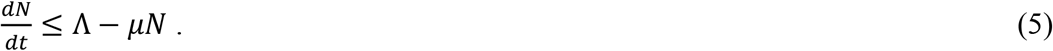

Solving equation (5) using Groonwall’s inequality gives 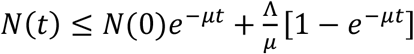. This means as 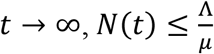 whenever 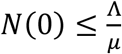. Hence, the non-negative solution set of the model equations (3) enters the feasible region, Ω, which is a positively invariant set.

### Positivity of the solutions

The following theorem proves that the solution of the model is non-negative for *t* ≥ 0.

Theorem 1. Let the initial solutions satisfy *S*(0)> 0, *E*(0)≥ 0, *Q*(0)≥ 0, *I*(0)≥ 0, *J*(0)≥ 0, *R* ≥ 0. The model has non-negative solutions which are contained in the feasible region, 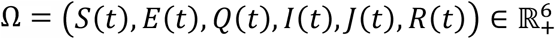.

Proof. From the first equation of (3),

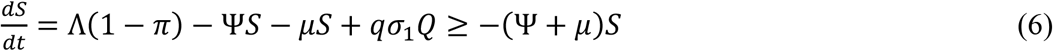

Solving (6) gives *S*(*t*) ≥ *S*(0)exp −(Ψ + *μ*) *t* ≥ 0.

In the same way,

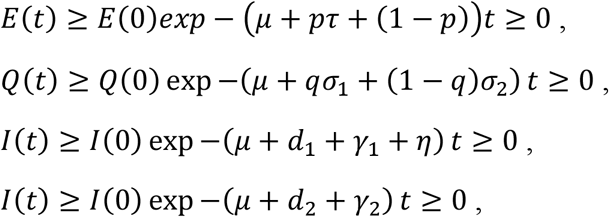

and

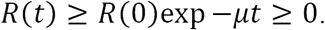

This shows that the solution set (*S*(*t*), *E*(*t*), *Q*(*t*), *I*(*t*), *J*(*t*), *R*(*t*))is non-negative for all *t* ≥ 0 since exponential functions and initial solutions are non-negative.

### Existence of disease-free equilibrium state

Let represent

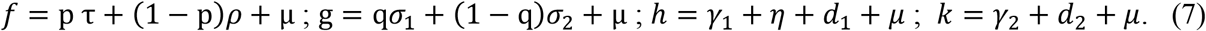

The model equations (3) becomes

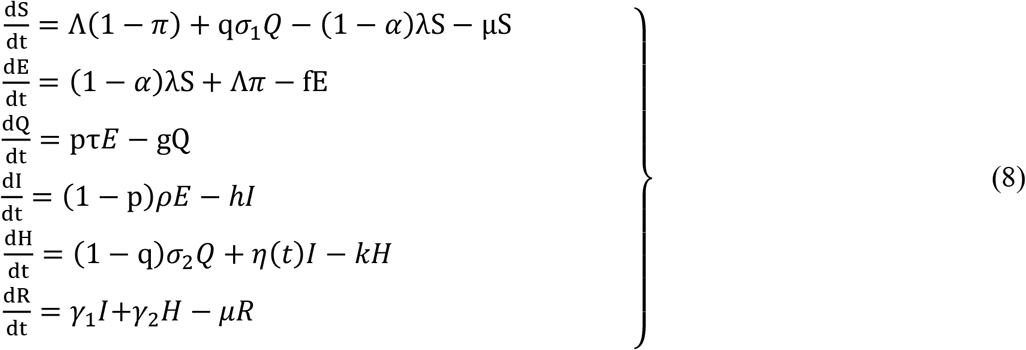

The disease-free equilibrium state, *E*_0_, is established when there is no infective immigrant into the population (i.e. *π* = 0).

The equilibrium state for the model equations (8) is at the state when 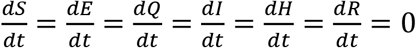 and this is solved simultaneously to give the disease-free equilibrium state. *E*_0_, as

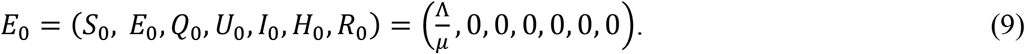

### Basic Reproduction Number

The basic reproduction number, *R*_0_, is a threshold quantity that predicts the spread of disease in the population. It is an average number an infective will infect people in a wholly susceptible population. If *R*_0_ < 1, the infection will die out. If *R*_0_ > 1, the infection will persist in the population. The approach of Next-generation method by Driessche and Watmough [28] is used to compute *R*_0_.

The rates of new infection and the transfer from in and out the infected compartments are given by

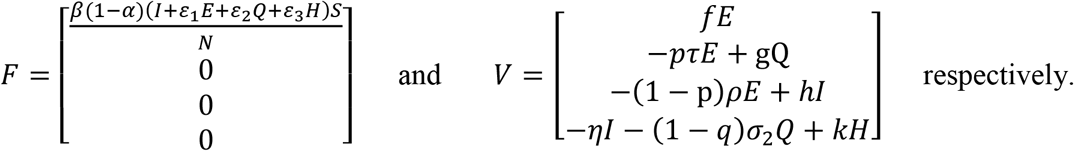

The partial derivatives of *F* and *V* at the DFE, *E*_0_, yield

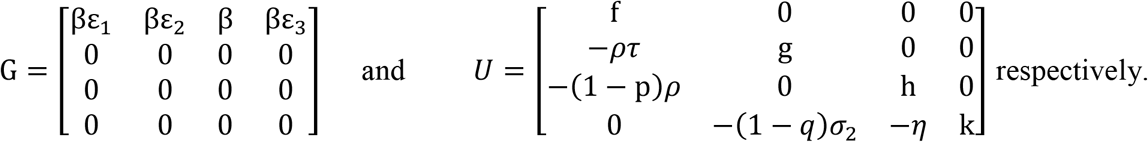

The basic reproduction number,*R*_0_, which is the spectral radius of the matrix, *GU*^*−*1^, given as

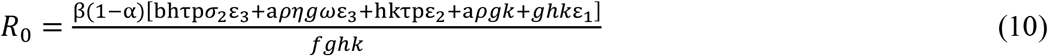

where

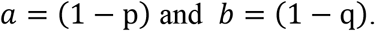

If p = 1, we have a perfect quarantine with no infectious not hospitalized individuals, i.e. *I* = 0. The reproduction number with perfect quarantine, *R*_0*q*_ is

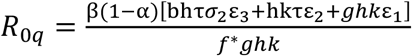

where

*f*\* = *τ* + *μ*

If *p* = 0, there is no exposed individual is quarantine, the basic reproduction number, *R*_0*p*_, is given by

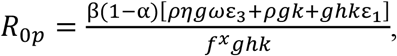

where

*f^x^* = *ρ* + *μ*.

### Existence of Endemic Equilibrium State

The equilibrium state for the COVID-19 model (8) is obtained by solving

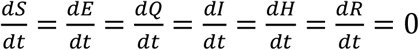

Simultaneously. This gives

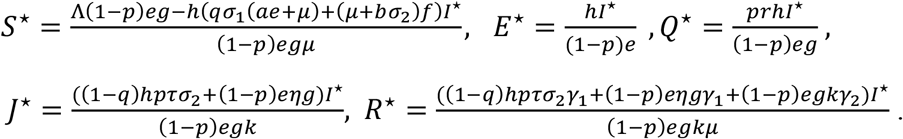

Substituting *S, E, Q, J, R* as *S*^⋆^, *E*^⋆^, *Q*^⋆^, *J*^⋆^, *R*^⋆^ and simplifying yields the following quadratic equation

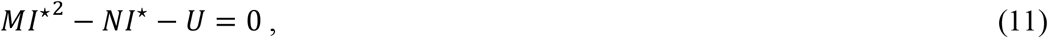

where

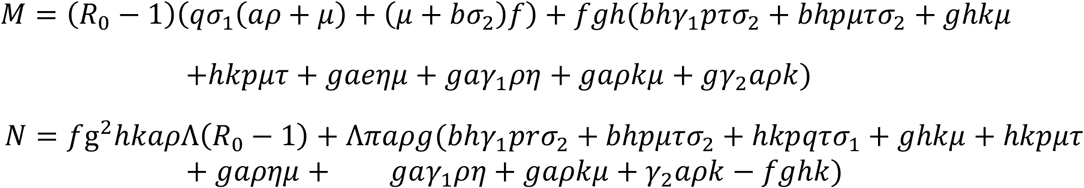

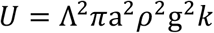

If *π* = 0, we have *U* = 0 and N = N^*^, in equation (11) where N^*^ = *f*g^2^*hkapΛ*(*R*_0_ *−* 1). So, equation (12) will become

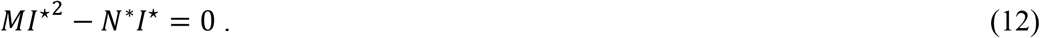

This implies from equation (12) when *π* = 0,

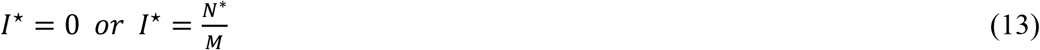

*I*^⋆^ = 0 corresponds to disease-free equilibrium (DFE) state, *E*_0_in equation (9), while 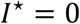 represents the endemic equilibrium state,*E*_1_ = (*S*^⋆^, *E*^⋆^, *Q*^⋆^, *I*^⋆^, *J*^⋆^, *R*^⋆^)when *π* = 0.

Substituting 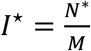 into *S*^⋆^, *E*^⋆^, *Q*^⋆^, *I*^⋆^, *J*^⋆^, *R*^⋆^gives the endemic equilibrium state, *E_1_*= (*S*^⋆^, *E*^⋆^, *Q*^⋆^, *I*^⋆^, *J*^⋆^, *R*^⋆^)where

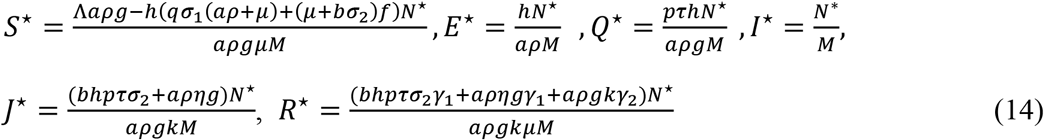

When *π* > 0, we have from equation (11) that

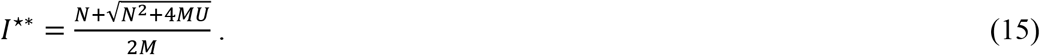

Substituting *I*^⋆*^ as *I*^⋆^ equation (15) into *S*^⋆^, *E*^⋆^, *Q*^⋆^, *I*^⋆^, *J*^⋆^, *R*^⋆^, we have

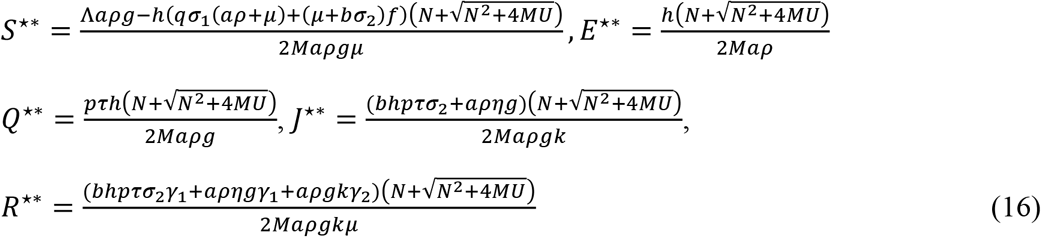

Equation (14) gives the endemic equilibrium state when *π* = 0 while equations (15) and (16) give the endemic equilibrium state when *π* > 0 provided that equations (15) and (16) satisfy the inequality,

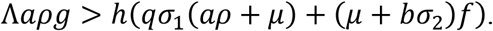

### Sensitivity Analysis of the model parameters

It is important to know the relative contribution of different model parameters responsible for the transmission and prevalence of any disease. This will help to identify where to focus interventions that will reduce the mortality and morbidity due to the disease. In this study, the sensitivity analysis is examined to identify crucial model parameters that will reduce the burden of the disease and also quantify the impact of each input parameter on the value of an outcome. The initial disease transmission is directly related to the basic reproduction number, *R*_0_. Therefore, we perform a sensitivity analysis on *R*_0_to identify the most critical parameters that will curtail the spread of COVID-19. We use forward normalized sensitivity index of *R*_0_ to measure the relative change in *R*_0_, to the relative change in the model parameter c. This is also defined using partial derivatives if *R*_0_ is a differentiable function of the model parameter, c, as is defined in Chitnis et al.[29]; Sanchez and Blowe [30] by

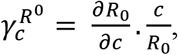

where, 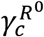 is the sensitivity index of *R*_0_of a parameter, c.

We compute the sensitivity indices for each parameter in *R*_0_. For instance, the sensitivity index of *R*_0_ for *β* is given as

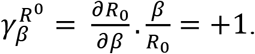

Table 2 shows the sensitivity indices of *R*_0_ for other parameters in *R*_0_ and their parameter values. The parameter values are taken from the literature on COVID-19, SARS and MERS.

**TABLE 2.** Parameter values and Sensitivity indices of *R*_0_ for the COVID-19 Model

| Parameter | Value | Index<br>Sign | Sensitivity Index Value | Source |
| --- | --- | --- | --- | --- |
| $\beta$ | 0.25 $day^{-1}$ | + | 1 | [31] |
| $\epsilon_1$ | 0.3 | + | 0.5386814 | [32] |
| $\epsilon_2$ | 0.1 | + | 0.1189200 | Assumed |
| $\epsilon_3$ | 0.1 | + | 0.0235887 | [32] |
| $e$ | 0.1429 $day^{-1}$ | + | 0.0062145 | [30] |
| $\sigma_2$ | 0.1259 $day^{-1}$ | − | 0.0704024 | [30] |
| $\alpha$ | [0,1] | − | 1 | Assumed |
| $\sigma_1$ | 0.07143 $day^{-1}$ | − | 0.0484410 | [30] |
| $d_1$ | 0.0079 $day^{-1}$ | − | 0.0149810 | [32] |
| $\gamma_1$ | 0.03521 $day^{-1}$ | − | 0.0667696 | [21] |
| $\gamma_2$ | 0.04255 $day^{-1}$ | − | 0.0020074 | [21] |
| $d_2$ | 0.0068 $day^{-1}$ | − | 0.0003209 | [32] |
| $\tau$ | 0.1259 $day^{-1}$ | − | 0.5386814 | [30] |
| $\eta$ | 0.13266 $day^{-1}$ | − | 0.2429498 | [30] |

From Table 2, the sensitivity indices with negative signs indicate that the value of *R*_0_ decreases when they are increasing while the sensitivity indices with positive signs show that the value of *R*_0_ increases when they are increasing. The sensitivity analysis shows that the most sensitive parameters are in the descending order of *β, α, ε*_1_, *τ, η*, and so on. These parameters will halt the spread of COVID-19 by reducing *β, ε*_1_, and increasing *a, τ, η*. It implies that the control parameters, *a, τ, η*, will reduce the spread of COVID-19 if they are increased. This is also shown in Figure 2 for the impact of *τ* and *η* on *R*_0_. This implies that increasing the rate of implementation of interventions such as awareness, quarantine and isolation in the exposed and infected not hospitalized population will halt the spread of COVID-19. In reducing *β, ε*_1_, we may consider the behavioral change in the transmission rate for further research.

**Figure 2.**
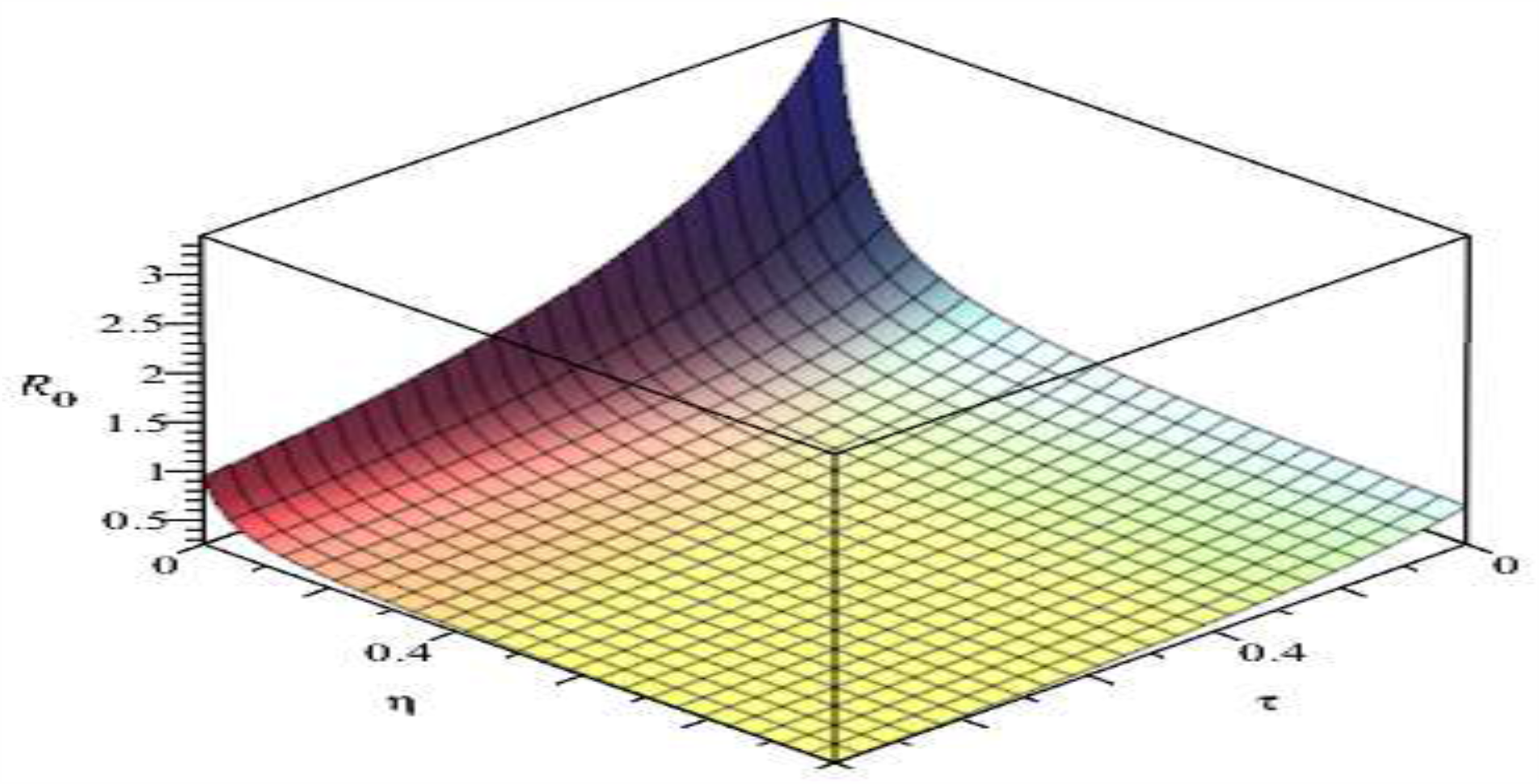
The effect of *η, τ*, on the basic reproduction number, *R*_0_. Here, other parameters are kept constant.

### Optimal control analysis

The control time-dependent parameters will be considered in this section. Our goal is to find an optimal control for public health education effort, *α*(*t*), quarantine effort for exposed individuals, *τ*(*t*), and isolation effort for infected individuals, *η*(*t*) such that

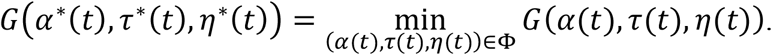

The necessary conditions that an optimal solution must satisfy are obtained by applying the Pontryagin’s maximum principle to the COVID-19 model of equation (3). This principle converts system (3) and equation (4) into a problem of minimizing pointwise Hamiltonian, H and is given as

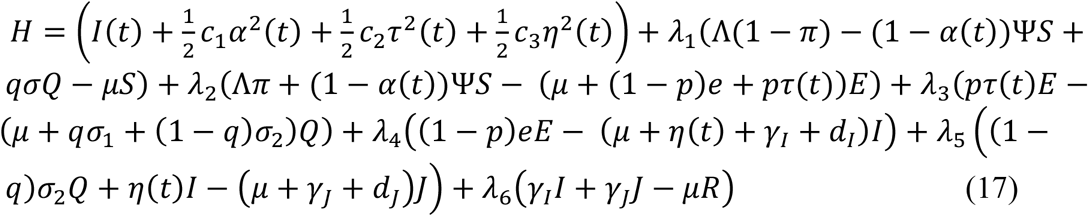

where *λ*_*i*_, *i* = 1, …, 6 denote the associated costate variables for the state variables *S, E, Q, I, J, R*. Using equation (17), we state the following theorem.

Theorem 2: Given an optimal control (*α*^*^(*t*), *τ*^*^(*t*), *η*^*^(*t*)) and solutions *S*^*°*^(*t*), *E*^*°*^(*t*), *Q*^*°*^(*t*), *I*^*°*^(*t*), *J*^*°*^(*t*), *R*^*°*^(*t*) of the corresponding state system (3) that minimizes *G*(*α*(*t*), *τ*(*t*), *η*(*t*)) over Φ, there exist costate variables *λ*_1_,*λ*_2_, *λ*_3_, *λ*_4_, *λ*_5_, *λ*_6_, that satisfy the following systems of equations

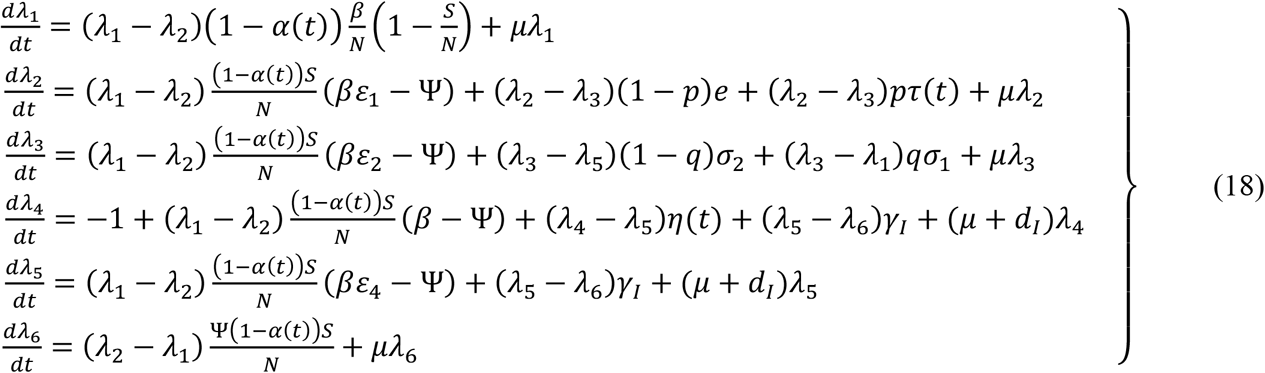

where Ψ is defined in equation (2) and final time conditions

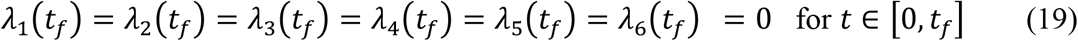

Also, the optimality conditions, *α*^*^(*t*), *τ*^*^(*t*), and *η*^*^(*t*) are given by

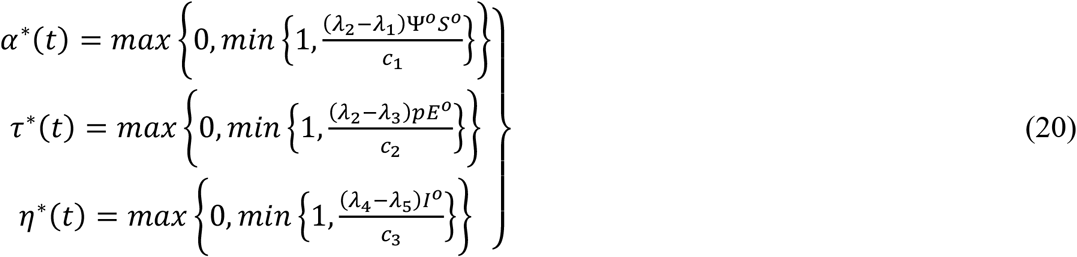

Proof:

Differentiating the Hamiltonian function, *H*, at the respective solutions of equation (3) and the optimal control with final time conditions, the differential equations governing the costate variables are obtained as follows

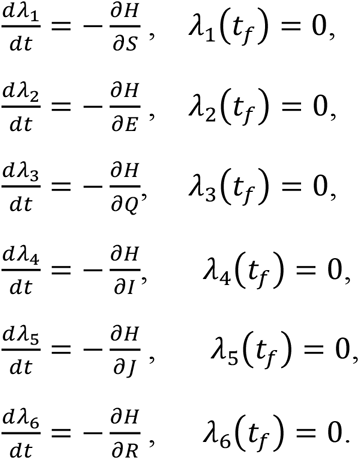

This gives the costate system in equation (18).

The optimality conditions are given in the interior of the control set Φ = {*α*(*t*), *τ*(*t*), *η*(*t*)|0 ≤ *α*(*t*), *τ*(*t*), *η*(*t*)≤ 1} as

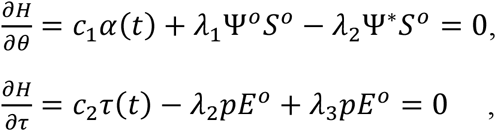

and

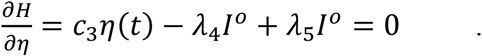

Solving for *α*(*t*) as *α*^*^(*t*), *τ*(*t*) as *τ*^*^(*t*), and *η*(*t*) as *η*^*^(*t*), yield

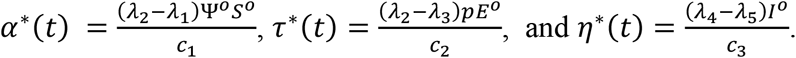

Thus, using the bounds of the controls, *α*^*^(*t*), *τ*^*^(*t*), and *η*^*^(*t*), the optimal control efforts in the compact form are given by equation (20).

The equations (3), (18) with optimality conditions (20) and the initial conditions,

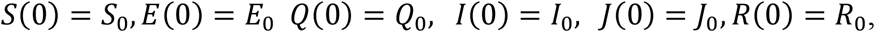

and final time conditions (19) gives the optimality system.

Owing to the priori boundedness of the state variables, the costate functions and the resulting Lipschitz structure of the ODEs, the uniqueness of the solutions of the optimality system is obtained for the small-time interval, *t*_*f*_. Hence the bounded solutions to the optimality system are unique for ∈’ [0, *t*_*f*_].

### Numerical Simulation

We carried out numerical simulations to investigate the impact of awareness, quarantine, isolation and the proportion of exposed individuals that will be quarantined. This is implemented using the parameter values and initial conditions from the literature on COVID-19, SARS and MERS [32, 33]. There are as follows, *S*(0)= 12 *Million, E*(0)= 1565, *Q*(0)= 800, *I*(0)= 695, *J*(0)= 326, *R*(0)= 200, *μ* = 0.000034, *Λ* = 600, c_1_ = 150, c_2_ = 300, c_3_ = 600. The other parameter values are given in Table 2. The optimality system is solved using the forward-backward sweep scheme. The details of the scheme are presented by Lenhart and Workman [34]. Many researchers have computed different values of the basic reproduction number for the person to person transmission, reservoir to person transmission, environmental transmission and some of their results have been compared with other types of coronaviruses, SARS and MERS and their results show almost similar results [5, 35]. Hence, we focus our numerical simulation on the impact of a different combination of control interventions with their different control profiles on the transmission dynamics of COVID-19.

## Discussion

A six compartmental model for the transmission dynamics of COVID-19 with quarantine, isolation and awareness as time-dependent control measures is examined using the work of Gumel *et al*. [14] as a guide. The model is for human-human transmission that involves imported cases and community spread. The model is proven to have an invariant region. This region is where the model is well-posed and makes biological sense to be carried out for the human population. The basic reproduction number, *R*_0_, is 1.51 when none of the exposed individuals are quarantined compared to when all of the exposed individuals are quarantined, *R*_0_ = 0.76. This means that a single infected person can transmit the infection to approximately two other persons when there is no quarantine while there is a possibility of stopping further transmission of infection when quarantine is implemented. However, there is a chance that some of the exposed individuals will evade quarantine due to fear of stigmatization and death. Therefore, public health education/awareness will help to correct their misconceptions and encourage them to accept the control measures. Furthermore, people that recovered need to share their experiences in quarantine and isolation centres with members of their community to enlist the cooperation of the entire community. When there is no isolation of the infected not hospitalized individuals in the population, the basic reproduction number, *R*_0_ = 2.5. This means that one infected not hospitalized person will infect approximately three persons in the population. The presence of isolation will help to reduce the number of infected not hospitalized individuals in the population. The simultaneous implementation of the three interventions reduces the number of infected individuals compared to the implementation of two interventions in the infected population (Figure 3a). This implementation takes about 64% input of awareness for 80 days, 58% input of quarantine for 95 days and 100% input of isolation for 98 days before they drop slowly to their lower bound (see Figure 4a). This does not achieve zero infection in the population which implies that more interventions are needed to eradicate the virus. On the other hand, the combined implementation of public health education and quarantine measures produces a better result for the exposed population (Figure 3b). It takes about 100% input of public health education for 90 days and 100% input of quarantine for 95 days to trace 2000 contacts in the exposed population (Figure 4b). It means that public health education/awareness should reach all the hooks and corners of the population. People need to be aware of the virus and also the creation of adequate awareness of COVID-19 among the population will facilitate contact tracing and quarantine of high-risk individuals. It will also help to identify those who do not qualify for quarantine but tested positive for COVID-19 to be isolated. This will reduce further transmission of COVID-19 in the population.

**Figure 3.**
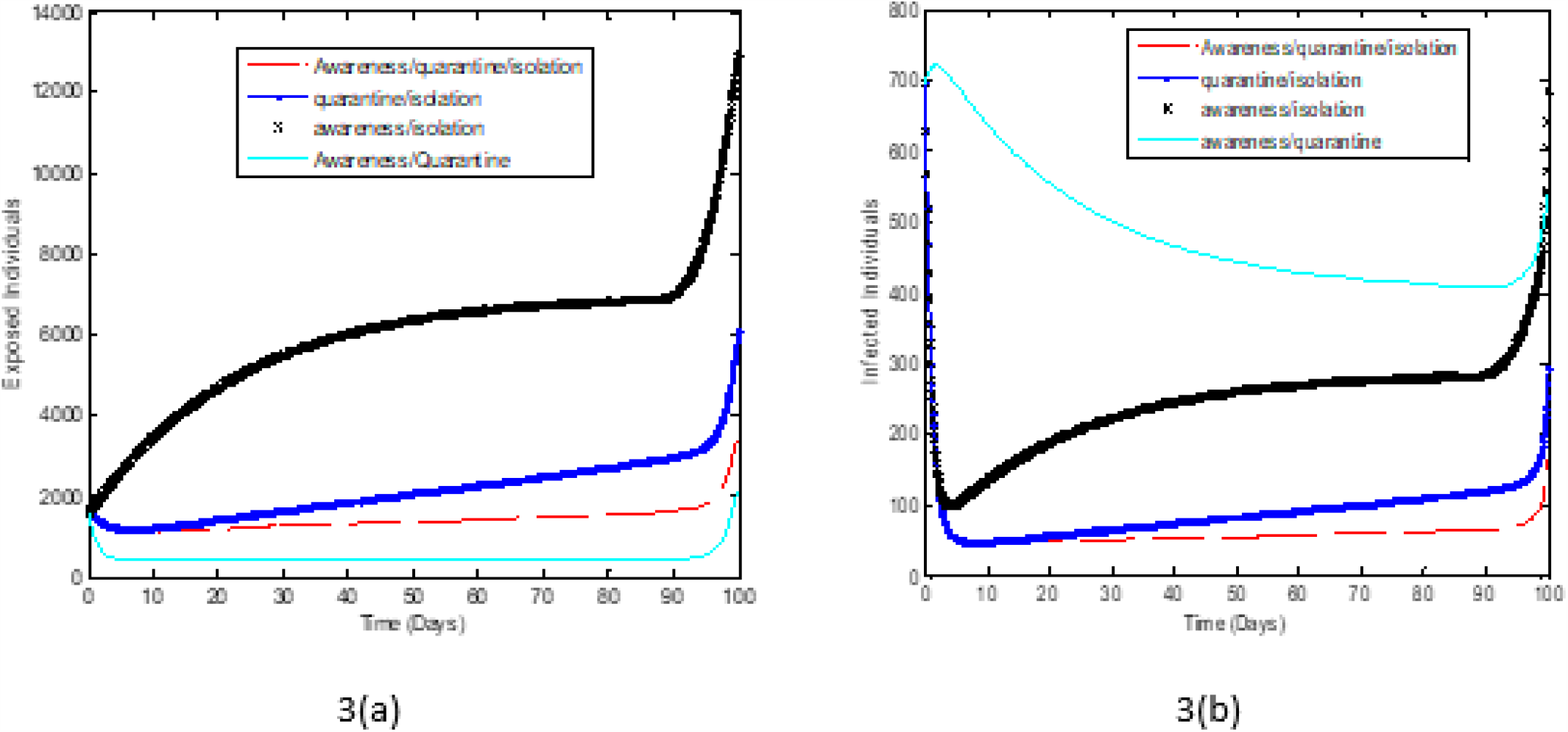
Optimal solutions for exposed individuals, (*E*), and infected individuals, (*I*), with different combinations of controls at a time.

**Figure 4.**
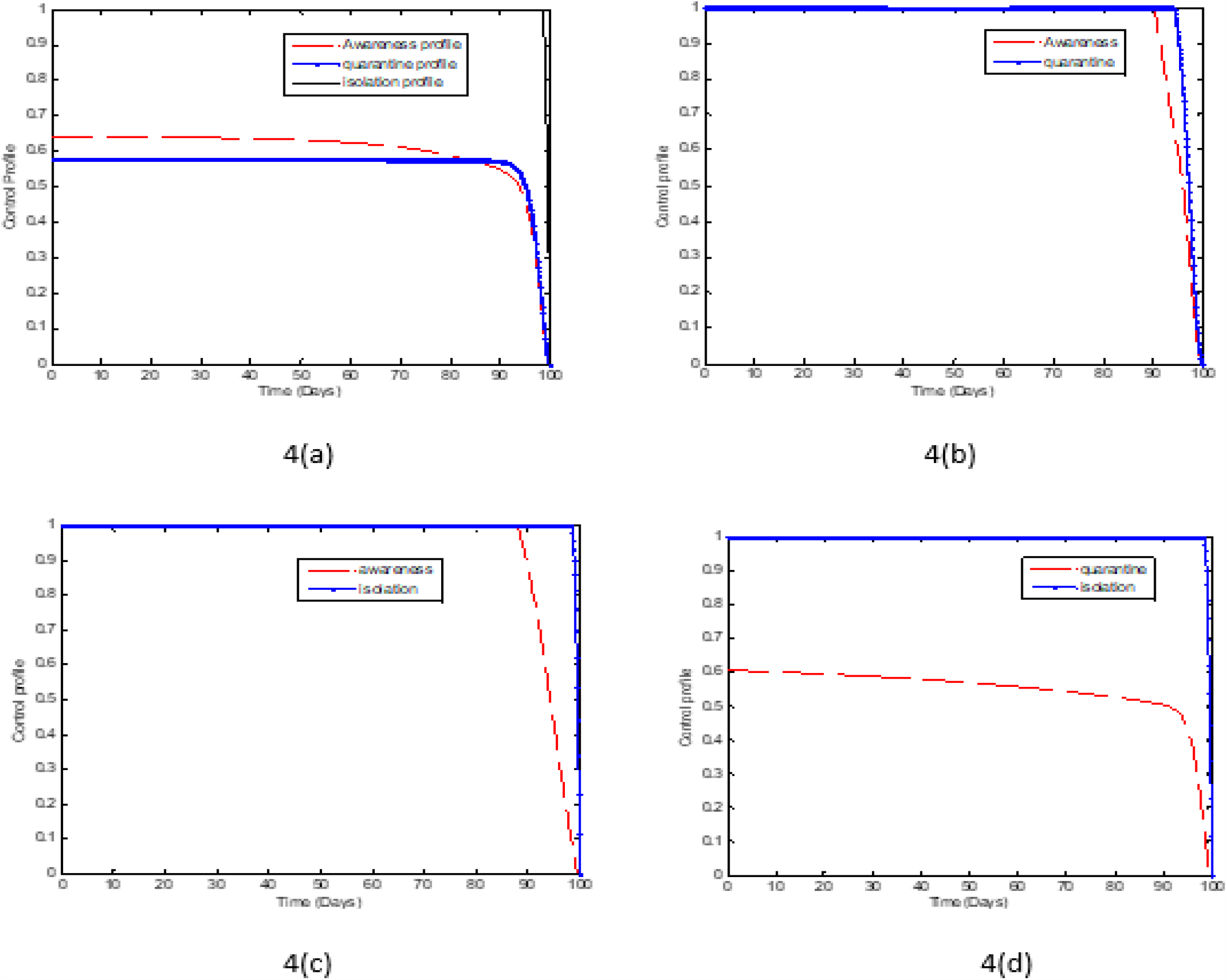
Optimal control profiles for different combinations of controls.

Furthermore, time is of importance in implementing these interventions (see Figure 5). It is observed that the time-dependent interventions reduce the number of exposed and infected individuals compared to the time-independent interventions. These interventions are good to implement early which is the first 2 -10 days of the outbreak. This will keep the burden of COVID-19 low. The virus will remain in the population for a prolonged time if there are no adequate interventions in place but it will eventually drop over time (see Figure 6). With the interventions such as public health education/awareness, quarantine and isolation, the number of exposed and infected individuals will reduce drastically within a short time but not to zero, leaving a residue of infected individuals with the potential to cause a further outbreak. This implies that COVID-19 will not be eradicated even with timely implementation of interventions unless a vaccine is developed.

**Figure 5.**
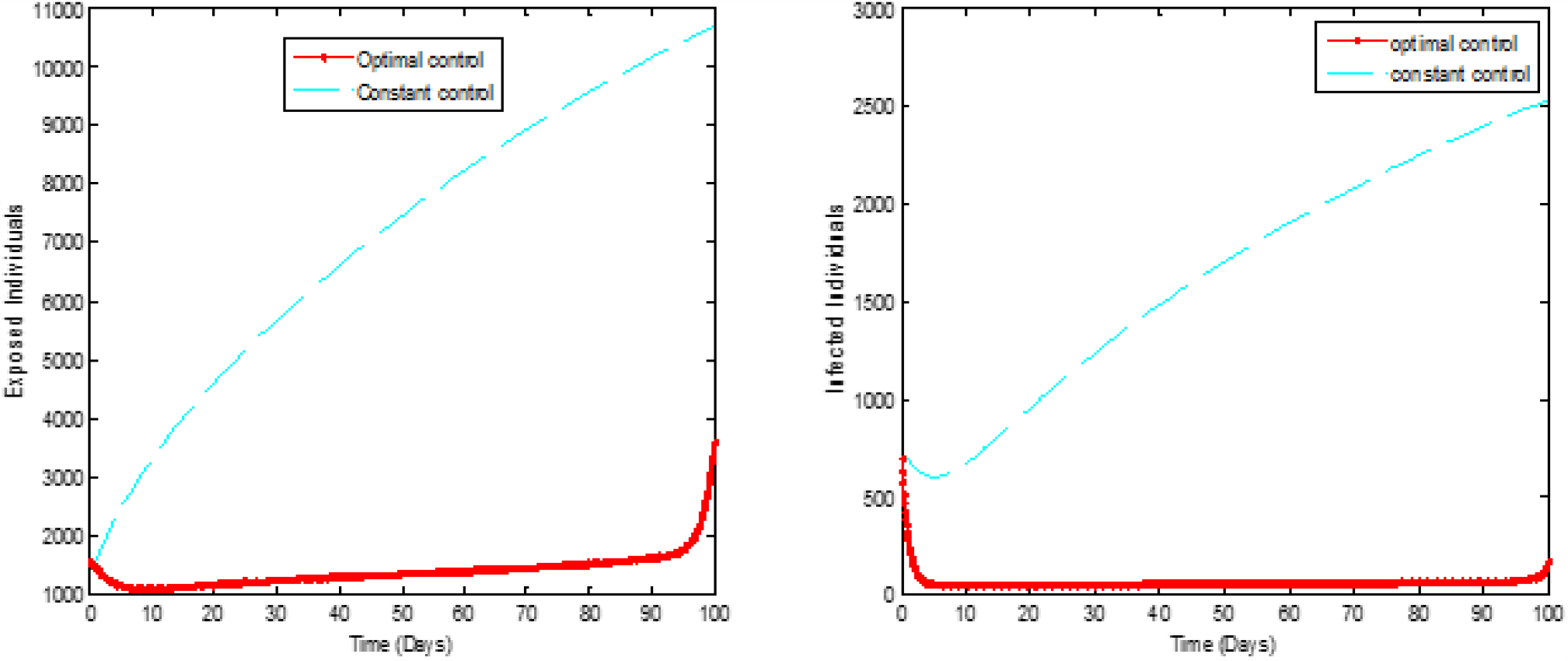
Optimal solutions and constant control for exposed individuals,(*E*), and infected individuals,(*I*).

**Figure 6.**
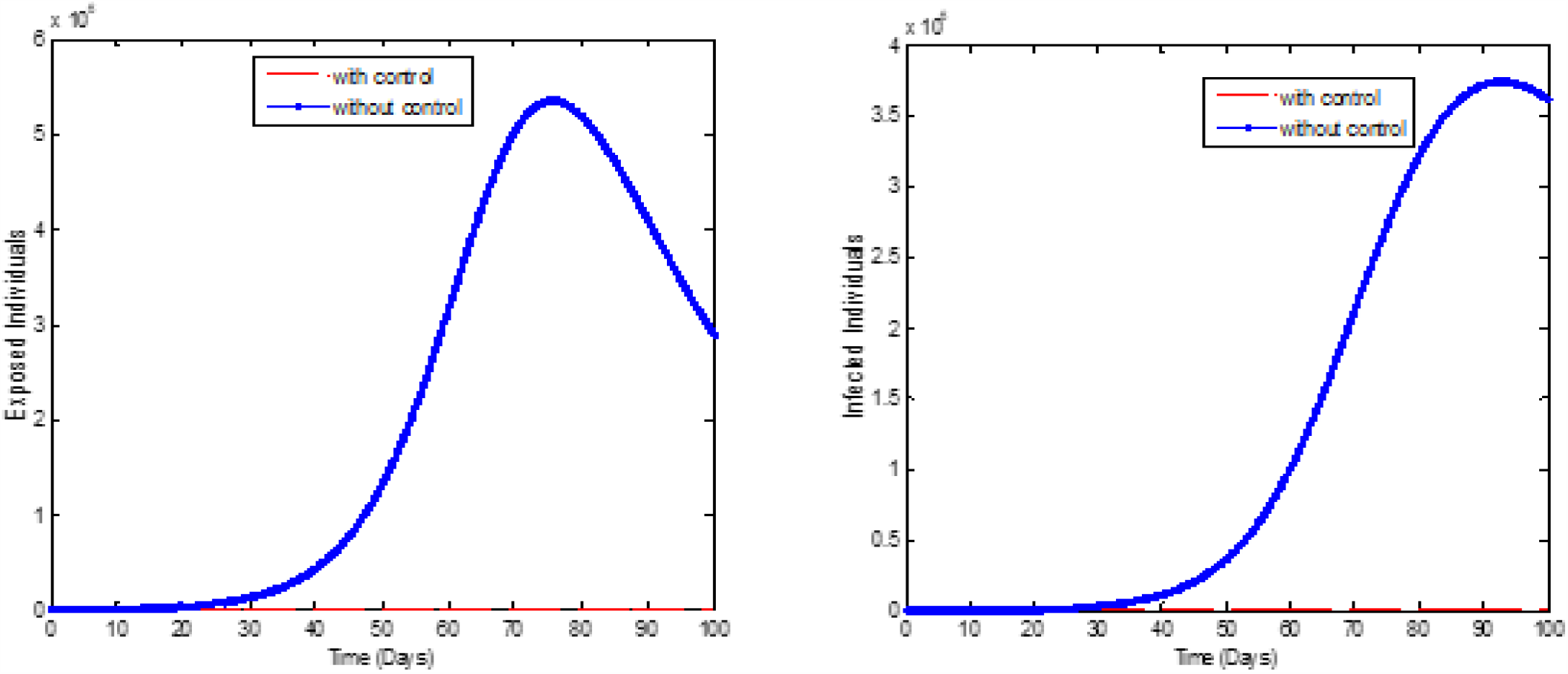
Optimal solutions with and without control for exposed individuals,(*E*), and infected individuals, (*I*).

When the proportion of the exposed individuals that are quarantine is increasing, it reduces the number of exposed individuals and infected individuals in the population (see Figure 7). When no exposed person is traced and quarantined in the population, the virus will remain in the population even when public health education/awareness and isolation interventions are present. When 30% of the exposed individuals are traced and quarantined immediately, the number of exposed individuals and infected individuals reduces to 2500 and 290 persons respectively. The number of exposed individuals and infected individuals is about 1200 and 90 persons when 50% of the exposed individuals are quarantined. Again, quarantining 80% of exposed individuals will result in about 700 exposed persons and 20 infected persons in the population. This does not eradicate the infection in the population. To achieve zero infection in the population, we postulate that, additional interventions such as mass testing, and vaccination need to be incorporated. These were not done in this work and would be the focus of further research. This is in line with the new directive by WHO for research.

**Figure 7.**
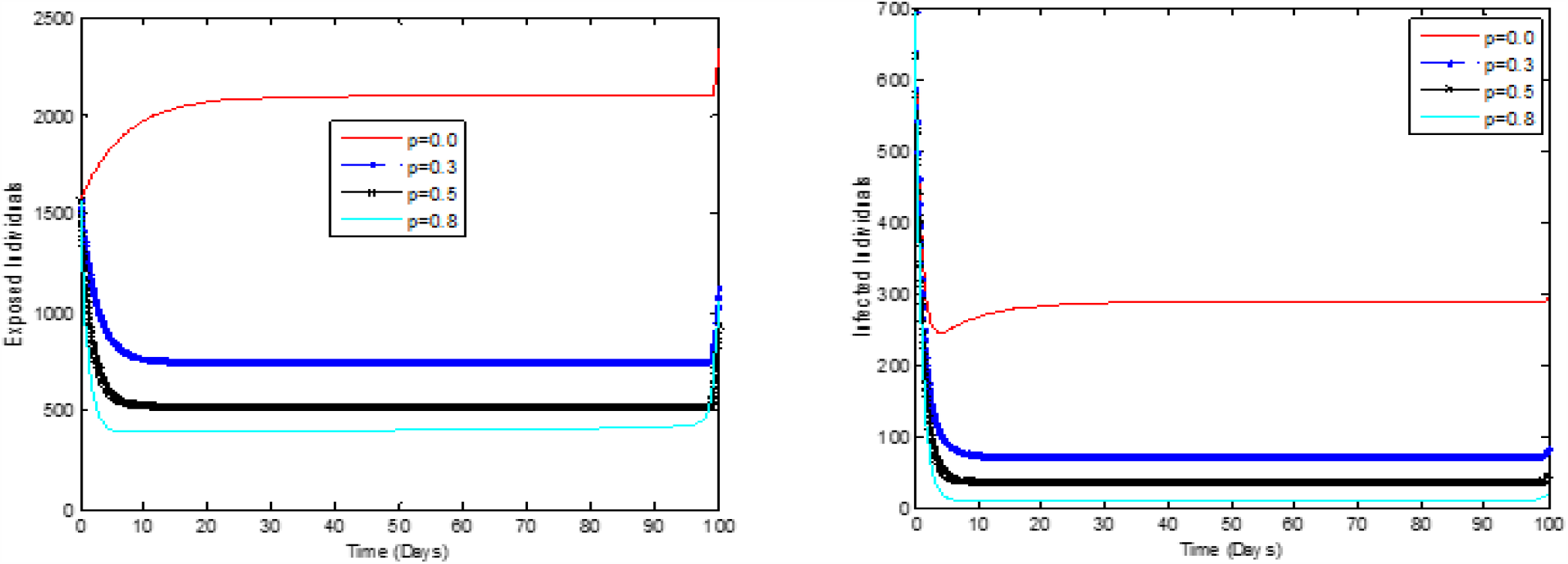
Optimal solutions for exposed individuals, (*E*), and infected individuals, (*I*), when more people are quarantine.

### Conclusion

In this paper, a new deterministic mathematical model of COVID-19 was formulated with quarantine, isolation and public health education as interventions. The model was also used as a prototyped to extensively investigate the contributions of these control measures to ascertain their individual and combined contributions in curbing the transmission and spread of COVID-19. The model analyses include the establishment of the Invariant region and positivity of the model, the existence of disease-free equilibrium, and computation of the basic reproduction number R0. It was found that the basic reproduction number, *R*_0_, is 1.51 when none of the exposed individuals are quarantined compared to when all of the exposed individuals are quarantined, *R*_0_ = 0.76. This means that a single infected person can transmit the infection to approximately two other persons when there is no quarantine while there is a possibility of stopping further transmission of infection when there is quarantine. It was also shown that when there is no isolation of the infected not hospitalized individuals in the population, the basic reproduction number, *R*_0_ = 2.5. This means that one infected not hospitalized person will infect approximately three persons in the population. The presence of isolation will help to reduce the number of infected not hospitalized individuals in the population. The simultaneous implementation of the three interventions reduces the number of infected individuals compared to the implementation of two interventions in the infected population. Furthermore, it was observed that the time-dependent interventions reduce the number of exposed and infected individuals compared to time-independent interventions. With the interventions such as quarantine, isolation and public health education, the number of exposed and infected individuals will reduce drastically within a short time but not to zero, leaving a residue of infected individuals with the potential to cause a further outbreak. This implies that COVID-19 will not be eradicated even with the timely implementation of these interventions.

Recommendation: Governments should ensure prompt implementation of quarantine, isolation and public health education in the COVID-19 fight in order to suppress its infectivity and mitigate the disease burden.

Further Research: Incorporation of mass testing and/or vaccination in the current model to ascertain its potential to eradicate COVID-19 in the population.

## Data Availability

The data used in the manuscript is cited within the manuscript.

## Data Availability

The data used in this article are included within.

## Acknowledgment

We want to appreciate Dr. Sunday Madubueze for proofreading the manuscript.

## References

[1] World Health Organization, “Coronavirus Disease 2019 (COVID-19), Situation Report -51, Data as reported by 11 March 2020,” Retrieve on 24/04/2020 from http://www.who.int/emergencies/disease/novel-coronavirus-2019/situation-reports.

[2] 1mg, “Coronavirus No-Panic Help guide,” retrieve on 24/04/2020,https://smef.org.uk/wp-content/uploads/2020/03/Corona-Ebook.pdf.pdf.pdf.

[3] B. Tang, N. L. Bragazzi, Q. Li, S. Tang, Y. Xiao and J. Wu, An Updated Estimation of the Risk of Transmission of the Novel Coronavirus (2019-nCov), Infectious Disease Modelling, 5: 248 – 255, 2020.

[4] W. G. Carlos, C. S. Dela-Cruz, B. Cao, S. Pasnick, and S. Jamil, “Novel Wuhan (2019-nCoV) Coronavirus,” American Journal of Respiratory and Critical Care Medicine, 2020.

[5] T. Chen, J. Rui, Q. Wang, Z. Zhao, J. Cui and L. Yin, “A Mathematical Model for Simulating the Phase-Based Transmissibility of a Novel Coronavirus,” Journal of Infectious Disease of Poverty, 9(24), 2020.

[6] S. He, S. Tang and L. Rong, “A Discrete Stochastic Model of the COVID-19 Outbreak: Forecast and Control,” AIMS Journal of Mathematical Biosciences and Engineering, 17(4): 2792 – 2804, 2020.

[7] P. Zhou, X. L. Yang, X. G. Wang, B. Hu, L. Zhang, W. Zhang et al., “A pneumonia Outbreak Associated with a New Coronavirus of Probable Bat Origin,” Nature, 579: 270 – 273, 2020.

[8] Q. Li, X. Guan, P. Wu, X. Wang, L. Zhou, Y. Tong et al., “Early Transmission Dynamics in Wuhan, China, of Novel Coronavirus-Infected Pneumonia,” the New England Journal of Medicine, 382(13): 1199 – 1207, 2020.

[9] N. Imai, A. Cori, I. Dorigatti, M. Beguelin, A. Donnelly, S. Riley and N. Ferguson, “Report 3: Transmissibility of 2019-nCoV,” Imperial College London (25-01-2020), 2020. https://doi.org/10.25561/77148.

[10] World Health Organization, “Coronavirus Disease 2019 (COVID-19), Situation Report -25” Data as reported by 14 February 2020. Retrieve on 24/04/2020 from http://www.who.int/emergencies/disease/novel-coronavirus-2019/situation-reports.

[11] Nigerian Centre for Disease Control (NCDC), “COVID-19 SITUATION REPORT: Situation Report 1 and Report 58,” Retrieve on 29/4/2020 from https://ncdc.gov.ng/disease/sitreps/?cat=14&name=An.

[12] P. Surico and A. Galeotti, “The Economics of a Pandemic: The Case of COVID-19: Lectures,” Wheeler Institute for Business and J-IDEA, Imperial College London Development, European Research Council, London Business School, 2020. https://icsb.org/theeconomicsofapandemic/.

[13] H. Lu, “Drug Treatment Options for the 2019-New Coronavirus (2019-nCoV),” Bioscience Trends, 14(1):69 – 71, 2020. https://doi.org/10.5582/bst.2020.01020.

[14] C. E. Madubueze, A. R. Kimbir and T. Aboiyar, “Global Stability of Ebola Virus Disease Model with Contact Tracing and Quarantine,” Appl. Appl. Math., 13(1):382 – 403, 2018.

[15] Y. Kim, S. Lee, C. Chu, S. Choe, S. Hong and Y. Shin, “The Characteristics Of Middle Eastern Respiratory Syndrome Coronavirus Transmission Dynamics in South Korea,” Osong Public Health and Research Perspectives, 7(1): 49 – 55, 2016.

[16] B. Gumel, S. Ruan, T. Day, J. Watmough, F. Brauer, P. Driessche van den, D. Gabrielson, C. Bowman, E. Alexander, S. Ardal, J. Wu and B. M. Sahai, “Modelling Strategies for Controlling SARS Outbreaks,” Proc. R. Soc. Lond. B, 271:2223-2232, 2004.

[17] J. T. Wu, K. Leung and G. M. Leung, “Nowcasting and Forecasting the Potential Domestic and International Spread of the 2019-nCoV Outbreak Originating in Wuhan, China: a modelling study,” THE LANCET, 395(10225): 689 – 697, 2020.

[18] S. Zhao, Q. Lin, J. Ran, S. S. Musa, G. Yang and W. Wang, “Preliminary Estimation of the Basic Reproduction Number of Novel Coronavirus (2019-nCoV) in China, from 2019 to 2020: A Data-Driven Analysis in the Early Phase of the Outbreak,” Int J Infect Dis, 92: 214 – 217, 2020.

[19] S. Zhao, S. S. Musa, Q. Lin, J. Ran, G. Yang and W. Wang, “Estimating the Unreported Number of novel Coronavirus (2019-nCoV) Cases in China in the First Half of January 2020: A Data-Driven Modelling Analysis of the Early Outbreak,” J Clin Med, 9(2), pii: E388, 2020.

[20] M. Shen, Z. Peng, Y. Xiao and L. Zhang, “Modelling the Epidemic Trend of the 2019 Novel Coronavirus Outbreak in China,” bioRxiv preprint first posted online, January 25, 2020.

[21] J. F. Rabajante, “Insights from early mathematical models of 2019-nCoV acute Respiratory disease (COVID-19) dynamics,” Preprint, February, 2020,

[22] R. Djidjou-Demasse, Y. Michalakis, M. Choisy, M. T. Sofonea and S. Alizona, “Optimal COVID-19 Epidemic Control Until Vaccine Deployment,” Preprint, April, 2020.

[23] S. E. Moore and E. Okyere, “Controlling the Transmission Dynamics of COVID-19,” Preprint, March, 2020. 2004.00443v2 [q-bio.PE] 2 April 2020.

[24] S. Edward and N. Nyerere, “A Mathematical Model for the Dynamics of Cholera with Control Measures,” Applied and Computational Mathematics, 4(2):53 – 63, 2015.

[25] S. Edward, E. M. Lusekelo, D. M. Ndidi and E. Simanjilo, “Mathematical Modelling of the Transmission Dynamics of Ebola Virus Disease with Control Strategies,” IJSBAR, 33(1):112 – 130, 2017.

[26] V. Kamyad, R. Akbarl, A. A. Heydari and A. Heydari, “Mathematical Modeling of Transmission Dynamics and Optimal Control of Vaccination and Treatment for Hepatitis B Virus,” Computational and Mathematical Methods in Medicine, Article ID 475451, 2014.

[27] S. F. Sadiq, M. A. Khan, S. Islam, G. Zaman, I. H. Jung and S. A. Khan, “Optimal Control of an Epidemic Model of Leptospirosis with Nonlinear Saturated Incidences,” Annual Research and Review in Biology, 4(3): 560 – 576, 2014.

[28] P. Van Den Driessche and J. Watmough, “Reproduction Numbers and Sub-Thresholds Endemic Equilibrium for Compartmental Models of Disease Transmission,” Mathematical Bioscience, 180: 29 – 48, 2002.

[29] N. Chitnis, J. M. Hyman and J. M. Cushing, “Determining Important Parameter in the Spread of Malaria through the Sensitivity Analysis of Mathematical Model,” Department of public health and epidemiology, 70: 1272–1296, 2008.

[30] M. A. Sanchez and S. M. Blowe, “Uncertainty and Sensitivity Analysis of the Basic Reproductive Rate: Tuberculosis As An Example,” Am. J. Epidemiol., 145: 1127–1137, 1997.

[31] J. Gjorgjiera, K. Smith, G. Chowell, F. Sanchez, J. Snyder and C. Castilo-Chavez, “The Role of Vaccination in the Control of SARS,” Mathematical Biosciences and Engineering, 2(4): 753 –769, 2005.

[32] X. Yan and Y. Zou, “Optimal and Sub-Optimal Quarantine and Isolation Control in SARS Epidemics,” Mathematical and Computer Modelling, 47: 235 – 245, 2008.

[33] N. Al-Asuoad, S. Alaswad, L. Rong and M. Shillor, “Mathematical Model and Simulations of MERS Outbreak: Predictions and Implications for Control Measures,” Biomath, 5(2016):1612141, 2016.

[34] S. Lenhart and J. T. Workman, “Optimal Control Applied to Biological Models,” Chapman and Hall/CRC, 2007.

[35] C. Yang and J. Wang, “A Mathematical Model for the Novel Coronavirus Epidemic in Wuhan, China,” MBE, 17(3):2708 – 2724, 2020.

